# Three-dimensional high-content imaging of unstained soft tissue with subcellular resolution using a laboratory-based multi-modal X-ray microscope

**DOI:** 10.1101/2025.07.13.25331466

**Authors:** Michela Esposito, Alberto Astolfo, Yang Zhou, Ian Buchanan, Alexei Teplov, Marco Endrizzi, Alexandra Egido Vinogradova, Olga Makarova, Ralu Divan, Cha-Mei Tang, Yukako Yagi, Peter D. Lee, Claire L. Walsh, Joseph D. Ferrara, Alessandro Olivo

## Abstract

With increasing interest in studying biological systems across spatial scales—from centimetres down to nanometres—histology continues to be the gold standard for tissue imaging at cellular resolution, providing an essential bridge between macroscopic and nanoscopic analysis. However, its inherently destructive and two-dimensional nature limits its ability to capture the full three-dimensional complexity of tissue architecture. Here we show that phase-contrast X-ray microscopy can enable three-dimensional virtual histology with subcellular resolution. This technique provides direct quantification of electron density without restrictive assumptions, allowing for direct characterisation of cellular nuclei in a standard laboratory setting. By combining high spatial resolution and soft tissue contrast, with automated segmentation of cell nuclei, we demonstrated virtual H&E staining using machine learning-based style transfer, yielding volumetric datasets compatible with existing histopathological analysis tools. Furthermore, by integrating electron density and the sensitivity to nanometric features of the dark field contrast channel, we achieve stain-free, high-content imaging capable of distinguishing nuclei and extracellular matrix.

## Introduction

Biomedicine is increasingly dependent on multi-modal and multi-scale imaging approaches to bridge vastly different length scales - from whole-organ imaging using standard clinical techniques, down to molecular level with *-omics*. Conventional histology remains the method of choice to connect macroscopic and nanoscopic domains, offering sub-cellular resolution and functional characterization of soft tissues through a diverse array of staining protocols. However, its inherently destructive nature and the introduction of tissue processing artefacts significantly limit its applicability to three-dimensional imaging [1]. Beyond histology, optical microscopy continues to be the gold standard for probing the microscale, but its efficacy quickly diminishes in thick tissue samples, due to the need for optical clearing [2].

The demand for maintaining subcellular resolution across increasingly thick samples has spurred growing interest in more penetrating probes, such as X-rays. Conventional X-ray imaging modalities, however, lack sufficient soft tissue contrast, making cellular structures difficult to resolve without staining, due to the weak contribution of the imaginary part (*β*) of the complex refractive index (*n* = 1 *− δ* + *iβ*) in the hard X-ray regime. Conversely, the unit decrement of the real part of the refractive index (*δ*) dominates at these energies and can be exploited by indirect measurement of the phase-shifts of waves travelling through the sample. The combination of X-rays’ penetrating power and the enhanced soft tissue contrast enabled by phase-sensitive techniques, combined with the penetrating power of X-rays, enabled significant breakthroughs with the use of synchrotron radiation [3, 4, 5, 6, 7, 8, 9, 10, 11].

Notably, synchrotron-based phase-contrast imaging has enabled a functional and multiscale investigation of the brain activity with subcellular resolution [12], the identification of pathological states of the brain (Alzheimer’s) at the cells’ nuclear level [13], a multi-modal and multi-scale visualisation of neurodegenerative state of the brain [14], visualisation of an entire mouse brain with sub-micron voxels [15], single cell imaging combined with confocal microscopy to identify inactive regions of the nuclear genetic material [16]. Whole organs can now be imaged with cellular resolution at synchrotron facilities, using a non-destructive hierarchical scan protocol [17] enabling multiscale imaging of a whole adult heart [18] and of a human placenta [19]. Crucially, Li et al. [20] showed that routine histochemical, immunohistochemical, DNA and RNA analysis are compatible with synchrotron imaging, suggesting the feasibility of linking morphology with the molecular domain for tissue samples.

The translation of phase-contrast imaging techniques from specialised synchrotron facilities to the laboratory environment, however, presents important challenges when subcellular resolution in unstained tissue is required. The potential of phase-contrast laboratory-based imaging at tissue scale has been shown in [21, 22, 23], enabling accurate identification of tumour margins, closely matching standard histological findings. However, these systems feature a resolution limit in order of 10-20 *µ*m, insufficient for cellular resolution. Multiple efforts have been reported that target cellular or subcellular resolution in tissue samples. Vagberg et al. [24] reported 3D imaging of unstained human coronary arteries and, while cellular resolution was claimed, only large (≈ 50 *µ*m) adipose cells or clusters of foam cells were visible. A comparative study of synchrotron and laboratory-based imaging of diseased human hearts is reported in [25], showing how the segmentation of the vascular network allows identification of different pathologies. Cellular resolution, however, is achieved only in the case of synchrotron imaging. Cellular resolution with a laboratory-based set-up has been demonstrated in [26] and [27] for renal tissue with eosin staining (a stain typically used in pathology), followed by critical point drying. The chosen tissue processing, however, limits possible downstream investigations (e.g. molecular analysis) and the choice of counterstains in histology. This strongly hinders the possibility of integrating this system in a multi-modal imaging workflow. Different tissue processing approaches were tested for brain imaging in [28], with samples scanned both with synchrotron radiation and with a laboratory-based CT scanner. While neurons and axons were visible with the laboratory-based system with all tissue preparation protocols, cells were only visible when samples had been stained with heavy metals (osmium tetroxide). Digital histology of unstained kidney glomeruli with an X-ray source has been reported in [29]. Although cellular-level imaging is claimed, the authors conclude that cell-like features reported in the article cannot be confirmed to be cells due to the lack of suitable verification (e.g. matching histology). Reichmann et al. [30, 31] report how synchrotron and laboratory-based phase-contrast imaging can enable the detection of pulmonary pathologies. However, cellular resolution was only achieved for synchrotron-based imaging. Cellular visualisation for brain tissue has been reported in [32, 33, 34]. Segmentation and a quantitative analysis of large and ramified Purkinje cells, as well as of smaller cells in the granular and molecular layers, are reported for both synchrotron and laboratory setup [33]. It has to be noted, however, that quantitative parameters for the segmented nuclei (e.g. electron density, radii, nearest neighbour distance) derived from laboratory-based measurements do not show a close agreement with the gold-standard synchrotron measurements. The laboratory-based imaging systems referenced above are all based on propagation-based approaches [35], providing access to a single, non-quantitative contrast channel. When propagation-based approaches are used, even in the synchrotron environment, electron density information extracted from tomographic scans rely on the restrictive hypothesis of sample homogeneity (i.e. constituted by at most two different materials) [36] and calibrated with respect to the substrate material of the sample [37], which requires an additional assumption on its chemical composition.

In this work we report the first demonstration of non-destructive fully quantitative three-dimensional imaging of unstained liver tissue, using a laboratory-based phase-contrast X-ray imaging system. Making use of quasi-monochromatic radiation, we ensure direct and quantitative access to electron density of the microscopy scale. Liver tissue, which typically features a high cell density, represents the optimal benchmark for demonstrating the capability of this system to detect the faint electron density difference needed to resolve cellular components. At the microscopic level, liver can also show pathological states such as vesicular fat metamorphosis and changes in the collagen arrangement in the extra-cellular matrix (ECM) leading to fibrosis. In this article, we show how the direct measurement of electron density allows the visualisation of hepatocytes nuclei over a mm^3^ of unstained Formalin-Fixed-Paraffin Embedded (FFPE) liver tissue, extracted from a conventional histology cassette. The achieved resolution (real 1 *µ*m, as opposed to voxel size typically quoted for commercial CT scanners) and contrast facilitated the automatic segmentation of cells’ nuclei and quantification of morphological parameters. The image quality of the measured electron density maps, combined with the accuracy of the segmentation workflow, enabled the use of a Machine Learning (ML) algorithm for converting measured three-dimensional electron density maps into three-dimensional H&E histology volumes.

The imaging system used in this work allows for the retrieval three complementary contrast channels [38]. In addition to phase (proportional to electron density), of particular interest is the dark field or Ultra-Small Angle X-ray Scattering (USAXS) that can identify the presence of ensembles of features below the system resolution [39], i.e. in the nanoscale. Here we demonstrate that, combining electron density and dark field maps, we can identify collagen fibrils in the ECM delivering high-content imaging.

## Results

### Three-dimensional cell visualisation

A 1-mm-diameter biopsy punch was extracted from FFPE liver tissue and imaged with the multi-modal X-ray microscope detailed in the Methods section. The design of the experimental workflow and a schematic representation of the imaging method are depicted in Figure 1(a). Figure 1 shows a three-dimensional rendering (b) and a virtual slice (d) of the electron density map of the punch biopsy. The orientation of the virtual slice was selected to closely match the orientation of an adjacent H&E slide obtained from the same sample (c). In addition to the original colour scale version (inset of panel (c)), histology was converted to grey scale for ease of comparison with the phase CT slice. Smaller Regions of Interest (ROI) from virtual and H&E slides (Figure 1 (e) and (f), respectively) reveal comparable features, including cell nuclei (hepatocytes, indicated by open arrows) and vesicular fat metamorphosis (filled arrows). The latter (also known as fatty change) is a pathological condition of the liver that, at the microscopic level, presents with the deposition of fat in the hepatocyte cytoplasm and the displacement of the hepatocyte nucleus at the periphery of the cell. ROIs for the virtual histology (Figure 1(g-j)) demonstrate the possibility to differentiate cell types based on their nuclear morphology. In addition to hepatocytes (open arrows), endothelial cells nuclei (notched arrows) and lymphocytes (arrowheads) are identified. For reference, the corresponding cell types are labelled in the histology of panel (f). Note that the colour scale of the electron density maps, here and elsewhere in the manuscript, was inverted for ease of comparison with histology, where nuclei appear darker on a brighter background.

**Figure 1.**
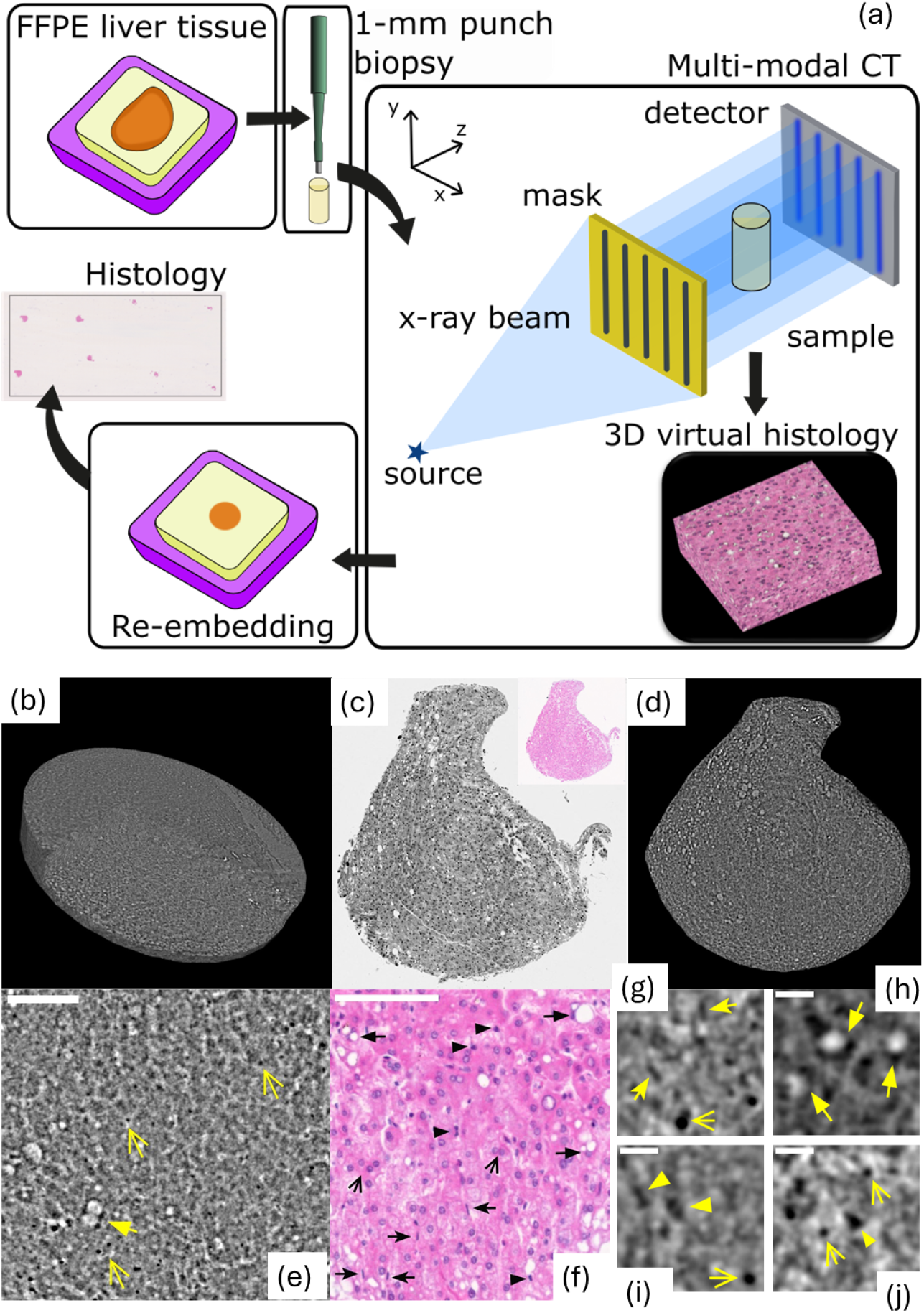
Three-dimensional imaging of the liver biopsy punch. (a) Schematic representation of the experimental workflow and diagram showing the imaging technique used in this work. (b) Volumetric representation of the electron density data. Sample diameter 1 mm. (c) An H&*E* histological slide converted to grey scale for ease of comparison alongside the original colour image in the inset. (d) A virtual slice of the electron density volume chosen at a similar orientation to the histology. Region of interest in the CT data (e) and histology (f). Scale bars 100 *µ*m. (g-j) ROIs extracted from the CT data (100 *×* 100 *µ*m^2^). Scale bar 20 *µ*m. Different arrow styles are used to highlight different histological features in panels (e-j). Open arrows are used to identify hepatocytes nuclei, notched arrows for endothelial cells nuclei, filled arrows for vesicular fat metamorphosis and arrowheads for lymphocytes nuclei. The colour scale of the electron density maps is inverted (darker = denser) in analogy with histology, to obtain darker nuclei on a brighter background.

### Segmentation and nuclear morphology

Using the automatic segmentation algorithm detailed in the Methods section, cell nuclei were segmented in the electron density CT data. Three-dimensional visualisations of segmented nuclei are shown in Figure 2(a-b) at different magnification. For each of the segmented nuclei, Signal-to-Noise ratio (SNR) and Contrast-to-Noise ratio (CNR) were calculated, comparing the signal measured in the nucleus and in the immediate surroundings (Figure 2 (c)). A mean SNR=18.33 ±0.05 and a mean CNR=1.673 ±0.003 suggests that nuclei can be easily detected above the background. The microscope used in this work allows for the quantitative retrieval of the phase signal, proportional to the electron density (see Methods section and Supplementary Materials). The electron density measured for segmented nuclei is reported in Figure 2 (d). The mean electron density measured for nuclei in liver tissue was *ρ*=314 *nm*^*−*3^, with standard deviation *σ*_*ρ*_=11 *nm*^*−*3^. This value is consistent with values reported in literature [13], albeit for a different tissue type. Measured parameters associated with nuclear morphology are shown in Figure 2, including nuclear volume (e) and maximum Feret diameter (f). The mean volume for the segmented nuclei was measured to be 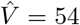 *µm*^3^ with a standard deviation of 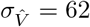 *µm*^3^, suggesting a strong heterogeneity in the nuclear size. The mean value of the three-dimensional Feret diameter was 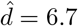 *µm*, characterised by a similarly broad distribution, resulting in a standard deviation of 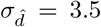 *µm*. Given that histology is inherently a two-dimensional technique while CT provides three-dimensional information, direct comparison of nuclear morphological properties between the two modalities is neither straightforward nor necessarily accurate. Nevertheless, the Feret diameter of nuclei measured in histological sections (see Methods section) yielded in a mean value of 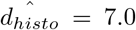 *µm*, which is comparable with the results obtained from our electron density CT. Additional morphological parameters, such as major (minor) axis length and eccentricity, are reported in the Supplementary Materials.

**Figure 2.**
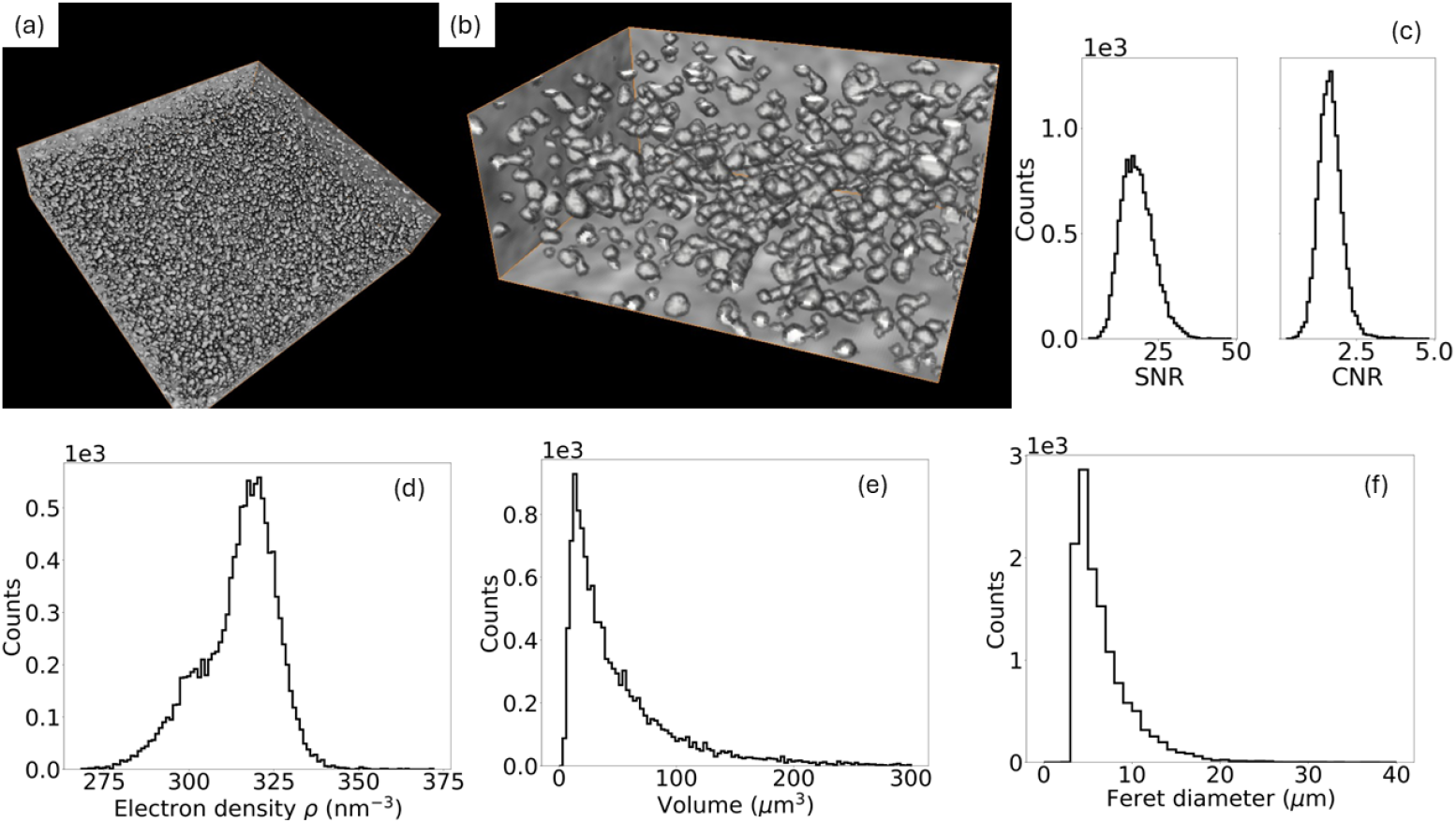
Automatic segmentation and nuclear morphology. (a) Three-dimensional rendering of segmented nuclei and a zoomed in region (b). (c) SNR and CNR distributions calculated for all segmented nuclei. Characterisation of nuclear morphology: electron density distribution (d), volume (e), and maximum Feret diameter (f).

### Virtual histology

While a wealth of quantitative information at cellular scale can be extracted from the dataset of Figures 1 and 2, direct interpretation of micro-CT images may be challenging for clinicians and biomedical researchers accustomed to conventional histological images. Furthermore, computational workflows designed for histological datasets may not be readily applicable to electron density maps. To address this limitation, we used a Machine Learning (ML)-based approach to convert electron density maps into a representation resembling H&E-stained histology. The style transfer was implemented using a Generative Adversarial Network (GAN), a class of models known for producing highly realistic synthetic images [40, 41] (see Supplementary materials). Figure 3 illustrates the outcome of the style transfer, with panels (a) and (b) highlighting the volumetric nature of the generated data. A two-dimensional comparison is also provided, showing paired images from both domains. Key liver features, such as hepatocyte nuclei and vesicular fat, are faithfully reproduced in the transformed images, demonstrating strong structural consistency between the modalities.

**Figure 3.**
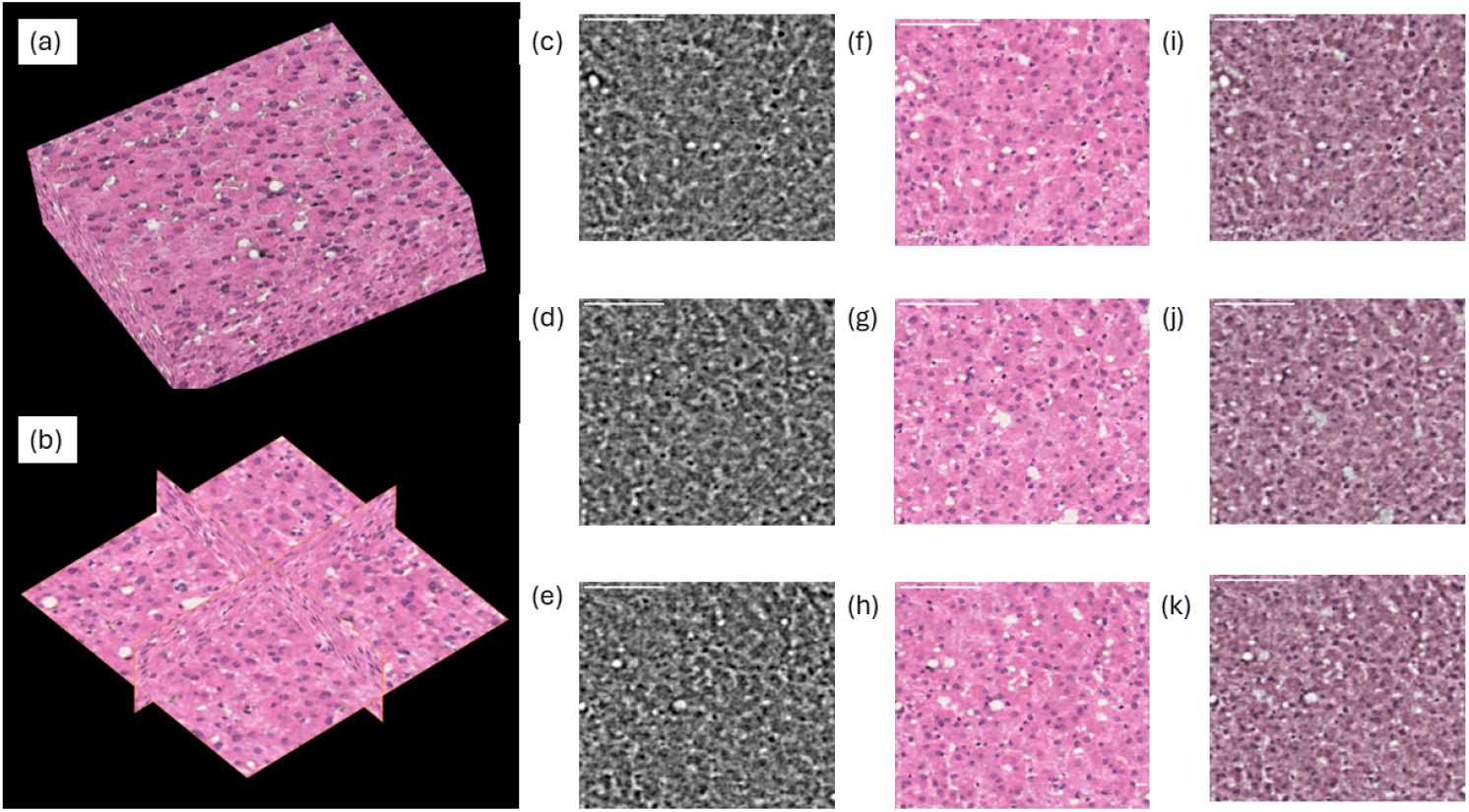
Three-dimensional virtual histology generated from the phase CT data. Panels (a) and (b) show a three-dimensional rendering of the generated histology data and cross-sectional views, respectively, highlighting the volumetric nature of the generated dataset. 2D slices (318 *×* 318 *µ*m^2^) are shown in panels (c)-(e) with correspondingly generated H&E histology (f)-(h). Each two pairs of images are overlaid and shown in panels (i)-(k) (50% transparency). Scale bars 100 *µ*m.

### High-content imaging

We demonstrate that the combination of complementary contrast channels, as provided by the multimodal X-ray microscope (described in detail in the Methods section), enables high-content imaging spanning both micro- and nanoscale length scales. Figure 4(a) shows a representative region of interest (ROI) from the sample, where phase contrast is rendered in greyscale and dark field contrast in shades of green. The two contrast channels reveal different structural features within the image. With a real resolution limit (as opposed to voxel size) of 1 *µ*m, the phase signal quantifies the electron density of sample features within the system’s spatial resolution, while the dark field channel highlights ensembles of sub-resolution features (i.e., at the nanoscale) [39]. A magnified region of the high-content image (Figure 4(c–e)) shows an individual cell, where the dark field signal is pronounced both inside the nucleus and at the cell periphery.

**Figure 4.**
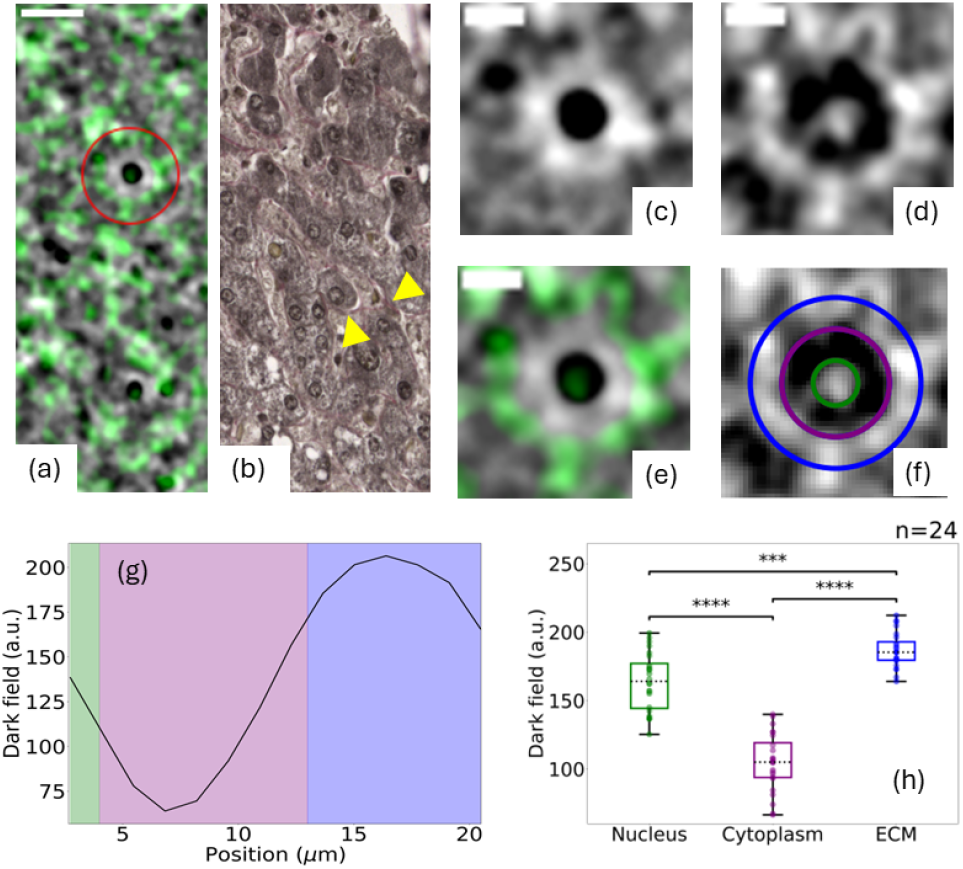
High-content histology: virtual staining using the dark field contrast channel. (a) High-content image of a ROI from the liver sample, with the phase signal displayed in greyscale and the dark field signal overlaid in green. Scale bar 25 *µ*m. (b) Trichome staining of a histological section from the same sample. Collagen in the extracellular matrix (ECM) appears pink and is marked with yellow arrowheads. (c-e) Zoom-in on an individual cell marked by the red circle in panel (a), displaying electron density map (c), dark field map (d), high-content image with phase in greyscale and dark field in greens (e). Three different regions can be identified in the dark field image, highlighted by green, purple and blue annuli in (f). Scale bars in (c-f) are 10 *µ*m. (g) Radial profile of the dark field signal in panel (f), with regions corresponding to the nucleus, cytoplasm, and ECM shaded in green, purple, and blue, respectively. (g) Values of the dark field signal associated with nucleus, cytoplasm, and ECM for individual cells (n=24). Boxplot elements: central mark indicates the median, and the bottom and top edges of the box indicate the 25th and 75th percentiles, respectively. The whiskers extend to the most extreme data points, not considered outliers. The statistical comparison was performed by Wilcoxon rank sum test (two-sided). A p-value*≤*0.05 is considered as statistically significant. We indicate p*≤*0.001 as (*\* * \**) and p*≤*0.0001 as (*\* * * \**).

Figure 4(g) presents a radial profile of the dark field signal for the cell shown in panel (d), centred on the nucleus. Two distinct peaks are observed. The first corresponds to the nucleus, as also evident in the electron density map (panel (c)). The second peak appears at the cell periphery, separated from the nuclear signal by a trough corresponding to the cytoplasmic region, which exhibits lower dark field intensity. An enhanced dark field signal associated with the nucleus has previously been reported [42], attributed to X-ray scattering from densely packed nuclear components. Similarly, the low signal in the cytoplasm is consistent with low density of scattering structures on the nanometric scale. The second peak at the periphery is likely associated with collagen in the extracellular matrix (ECM), known to produce strong X-ray scattering due to its microscopically heterogeneous structure [43, 39]. The three regions—nucleus, cytoplasm, and ECM—are shaded green, purple, and blue, respectively, in the radial profile (panel (g)) and are annotated with correspondingly coloured annuli in the dark field image (panel (f)). A histological section from the same tissue, stained with trichrome (a collagen-specific stain), is shown in Figure 4(b), where collagen adjacent to the cell membrane is highlighted by yellow arrowheads.

Finally, Figure 4(h) presents a boxplot of the dark field signal measured across 24 individual cells, for each of the three identified regions. The plot reveals statistically significant differences in signal intensity between the three regions (p *≤* 0.001 and p*≤* 0.0001), further supporting the ability of the dark field channel to observe nanoscale structural domains.

## Discussion

The first key result presented in this article is the demonstration that multi-modal X-ray microscopy enables three-dimensional histology with subcellular resolution (Figure 1), offering a viable alternative to conventional histology in bridging the gap between macroscale and nanoscale imaging of soft tissues. Conventional histology, making use of specialist stains (with H&E being the most common choice) produces intrinsically segmented images. In a H&E histology slide, nuclei appear in purple/blue and cytoplasm and extracellular matrix in pink. Hence, it is key for the successful translation of the technology presented in this work to include a robust segmentation stage. The spatial resolution and image contrast achieved with the X-ray microscope allow for the detection of nuclei with a high SNR (see Figure 1 (c)), well above the Rose criterion of SNR=5. This enabled the straightforward implementation of an automatic segmentation workflow (see Supplementary Materials). The simplicity and speed of the algorithm, combined with the lack of need for external inputs (e.g. seeding, training datasets), makes it possible to envision automatic scanning and segmentation workflows facilitating high-throughput imaging.

Leveraging on the reliable segmentation of cells’ nuclei and the availability of ML algorithms for style transfer, we demonstrated three-dimensional X-ray virtual histology with H&E staining. The resulting datasets are fully compatible with existing histopathological analysis tools and are immediately interpretable by clinicians and life scientists trained in traditional histology. This result represents a key advancement for volumetric tissue imaging. In fact, conventional histology can lead to misinterpretation of three-dimensional structure as a result of distortions from processing and sectioning [44]. While three-dimensional reconstruction of multiple serial slides is possible, this is often problematic as it leads to the exhaustion of limited available tissue. Additionally, the reliability of the reconstructed volumes is often limited, as biases can be introduced in the reconstruction process [1]. Here we demonstrated that the method proposed in this paper can overcome these limitations, by providing inherently three-dimensional H&E volumes in a non-destructive fashion, and so preserving tissue for ancillary investigations.

Another core strength of this approach is the quantitative nature of the intensity-modulation phasecontrast technique enabled by the use of quasi-monochromatic radiation, which provides direct measurement of electron density. Unlike propagation-based techniques [33], which rely on restrictive assumptions on sample homogeneity and chemical composition to obtain an estimate of electron density [37], the proposed method measures electron density directly, with the added advantage of a potential lower uncertainty and is less prone to systematic errors. By using nanoscale resolution imaging techniques at specialised synchrotron facilities, Bhartiya et al. [45] demonstrated that quantifying nuclear electron density can provide crucial information on the nuclei’s genetic components and give information on cells’ health and on chromosomal aberrations. Our work makes this possible in the standard laboratory environment, combining cell type identification using nuclear morphology (e.g. volume, Feret diameter, eccentricity, see Figure 1 (f-i) and Supplementary Materials) with quantification of electron density.

The second major innovation of this work is the possibility to produce stain-free high-content imaging, combining complementary contrast channels. The use of different histological stains is common practice in histology, as it allows for identifying different functional structures, e.g. in this work we showed H&E and trichrome stains to highlight nuclei and collagen in the ECM (Figure 1 and 4). A main limitation associated with the use of multiple stains, beyond cost and time requirements, is that the process of obtaining histological slides is destructive, resulting in the impossibility of running comparative analysis on the same tissue slice. Here we showed that combining electron density and dark field maps allowed the stain-free identification of nuclei and ECM, making this method a high-content imaging modality.

The X-ray microscope developed in this work is a pre-commercial prototype designed for imaging tissue volumes on the order of 1-mm^3^. Nevertheless, its design allows for scalability to larger volumes, enabled by its aperture-driven resolution [46], which arises from the use of intensity-modulation masks. In contrast, propagation-based systems aimed at virtual histology with cellular resolution typically have their spatial resolution constrained by the chosen detector pixel size and source focal spot size. For this reason, nano-focal sources with high geometrical magnification [26, 27] or micro-focal sources with optical magnification [33, 29] were used. Since spatial resolution is intrinsically linked to the source and detector characteristics, scaling to accommodate larger sample sizes remains a significant challenge. This often results in reduced resolution when imaging at larger fields of view (FoVs). In the prototype developed here, however, spatial resolution is decoupled from these parameters, allowing for imaging of larger FoVs without sacrificing resolution.

## Methods

### Liver tissue

Formalin-Fixed and Paraffin-Embedded (FFPE) liver tissue, mounted on a conventional histology cassette, was obtained from the Department of Pathology at the Memorial Sloan Kettering Center^1^. A 1-mm diameter biopsy punch was obtained from the paraffin block and imaged with the X-ray microscope. See Figure 1(a) for a schematic representation of the experimental workflow. The biopsy core was then re-embedded in paraffin, sectioned in 4 *µ*m-thick slices, stained with hematoxylin and eosin (H&E), and then scanned with a Whole Slide Imaging Scanner (WSI) system (Nanozoomer S60, Hamamatsu Photonics, Hamamatsu, Japan) at 40 *×* magnification (0.23 *µ*m/pixel). An additional slice was stained with Trichrome and scanned using the same WSI equipment. It should be noted that the obtained histological images show a *≈*10% increase in the longitudinal direction compared to CT, due to shear deformations as a result of the slicing process. Nuclei in the H&E histology images were segmented and their size quantified using the function *Cell detection* of the commercially available software QuPath (v. 0.5.1) [47].

### X-ray microscope

The X-ray microscope used in this work consists of a rotating anode Cu source (Rigaku) with a focal spot size of 70 *µ*m. The X-ray beam passes through a flat multi-layer monochromator [48] selecting the Cu K_*α*_ lines ( *≈*8 keV). An intensity-modulation mask is placed at 70 cm from the focal spot. The mask, detailed in [49], is a 10-*µ*m-thick Au membrane featuring 1 *µ*m wide apertures on a 7.5 *µ*m period. The sample, whose position is adjusted with six degrees of freedom, is placed in close proximity to the mask. A propagation distance of 35 mm is set between sample and detector. The detector (Rigaku, XSight™ Micron LC X-ray sCMOS), based on an indirect detection approach, features an effective pixel size of 0.65 *µ*m.

### Intensity-modulation phase-contrast imaging

The X-ray microscope is based on the use of an intensity-modulation mask to detect phase shifts. The mask shapes a broad X-ray beam into an array of beamlets so that changes in intensity, position, and width of each beamlet can be quantified and associated to the sample’s properties [50]. A schematic diagram of the intensity-modulation approach is shown in Figure 1(a). Assuming a bell-shaped distribution for the shaped beamlets, they can be individually characterised in terms of amplitude *A*, central position *µ* and variance *σ*^2^. Measuring these parameters with {*A, µ, σ}* and without {*A*_0_, *µ*_0_, *σ*_0_} sample at each angular position *θ* of a tomographic scan, allows the retrieval of transmission *L*, refraction *R* and dark field *DF*. For a monochromatic X-ray beam and following [51], these can be written as:

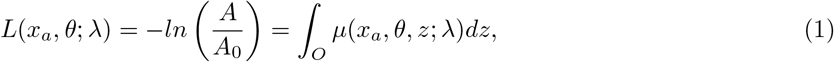

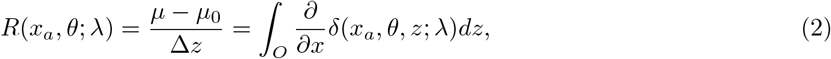

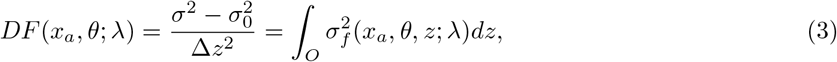

with Δ*z* being the propagation distance between sample and detector along the optical axis *z, x*_*a*_ the location of a specific mask aperture (and hence beamlet), *λ* the wavelength of the monochromatic beam, *O* the extent of the sample along the optical axis, and *µ* = (4*π/λ*)*β* the linear attenuation coefficient. *β* and *δ* are the imaginary part and the decrement from unity of the real part of the complex refractive index, respectively. 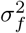 is the variance of the scattering distribution function *ϕ*. Equations 1-3 can be inverted by tomographic reconstruction to obtain quantitative three-dimensional maps of *µ, δ* and 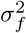. Of particular interest for this work is the phase signal *δ*, as it allows obtaining maps of *δ* that can be directly and quantitatively linked to the electron density *ρ* By

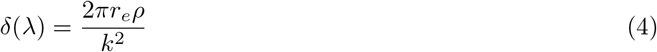

with *r*_*e*_ the classical electron radius and *k* = 2*π/λ* the wave number. The quantitativeness of the retrieved electron density is discussed in the Supplementary Materials.

### Imaging, retrieval and reconstruction

The biopsy punch was imaged using the X-ray microscope. Projection images were obtained over 360^*°*^ in steps of 0.25^*°*^. At each angular position, the sample was *dithered*, i.e. stepped in sub-aperture steps (0.75 *µ*m) for a total shift equal to the mask period (7.5 *µ*m) to deliver full sample illumination [52]. An exposure time of 30 s per frame was used. Options for reducing the dithering step process are discussed in the Supplementary Materials. Retrieval of transmission *L*, refraction *R* and dark field *DF* was performed at each angular and dithering position using Equations 1-3, and the resulting images for each dithering step interleaved, to yield images with resolution corresponding to the size of the mask apertures (1 *µ*m). Refraction images (see Equation 2) were integrated along the direction of phase sensitivity (*x*) using the Hilbert transform. Each of the three independent tomographic datasets was reconstructed using the Core Imaging Library (CIL) [53, 54]. Tomographic reconstructions were performed using a non-iterative algorithm based on the Tikhonov regularisation, with the gradient operator as regularisation operator. The optimisation problem solved using the Conjugate Gradient Least Square (CGLS) method. Additional computations tasks, including segmentation and style transfer, are detailed in the Supplementary Materials.

## Data Availability

All data produced in the present study are available upon reasonable request to the authors

## Acknowledgments

Research reported in this publication was supported by the National Institute of Biomedical Imaging and Bioengineering of the National Institutes of Health under Award Number R01EB028829. The content is solely the responsibility of the authors and does not necessarily represent the official views of the National Institutes of Health. Mask fabrication performed at the Center for Nanoscale Materials, a U.S. Department of Energy Office of Science User Facility, was supported by the U.S. DOE, Office of Basic Energy Sciences, under Contract No. DE-AC02-06CH11357. AO was supported by the Royal Academy of Engineering under the “Chairs in Emerging Technologies” scheme (CiET1819/2/78) nd PDL via (CiET1819/10). YZ/PDL/CLW were supported in part by CZI grant DAF2022-316777.

## Footnotes

1 The paraffin block was obtained from discarded tissue and, as such, exempt from ethical approval of Internal Review Board

## Notes

### Competing Interest Statement

JDF is a Rigaku employee and AEV was a Rigaku intern at the time this research was carried out; Rigaku has a potential interest in the commercial exploitation of the results presented in this article. OM and C-MT are Creatv Microtech employees; Creatv Microtech fabricated the X-ray masks used in this work and has a potential interest in their commercialization. MEn and AO are named inventors on patents held by UCL protecting some of the imaging technology discussed in this paper.

### Author Declarations

The IRB of Memorial Sloan Kettering Cancer Center( New York, USA) waived ethical approval for this work as it only used discarded pathology tissue

