## Supplementary material for "Three-dimensional high-content imaging of unstained soft tissue with subcellular resolution using a laboratory-based multi-modal X-ray microscope"

#### Automatic segmentation of electron density maps

Electron density maps, obtained from the phase CT using Equations 2 and 4 of the main manuscript, were segmented to isolate individual cell nuclei. An example electron density image is shown in Figure 1 (a) with inverted colour scale to match grey scale histology (Figure 1 (c) of the main manuscript). Black and white top hat filters were applied to the CT datasets to identify nuclear and non-nuclear regions, respectively. The result of applying top hat filters to the electron density map is shown in Figure 1 (b). Top hat filters were used as labels for a random-walk-based segmentation algorithm [1]. Figure 1 (c) shows the outcome of the random walk algorithm, with nuclei contours highlighted in green.

Segmentation labels were subsequently post-processed to cluster connected voxels, i.e. individual nuclei. The function *label* from the *measure* package in Scikit-Image (v. 0.22) library was used, using 1-connectivity. Properties of each labelled nucleus were then extracted using the function *regionprops* from the same library, including average intensity (i.e. electron density), volume, Feret diameter and the lengths of the major and minor axes. Eccentricity was calculated as the ratio of the latter two parameters. Labels were then used for calculating Signal-to-Noise and Contrast-to-Noise ratio (SNR and CNR) for individual nuclei. The mean signal $\bar{S}_i$ was calculated across all voxels assigned to each nucleus $i$. The mean background $\bar{B}_i$ was calculated as the mean voxel intensity within the bounding box of each segmented nucleus $i$, expanded by 2 voxels in each direction, while excluding pixels belonging to the nucleus itself. This process ensures that the signal in each nucleus is compared to the background in its immediate vicinity. SNR and CNR were calculate as follows: $SNR = S_i/\sigma_{B,i}$ and $CNR = S_i - B_i/\sigma_{B,i}$, with $\sigma_{B,i}$ being the standard deviation of the background signal of the *i-th* nucleus ($B_i$).

#### Validation of the segmentation workflow

In the main manuscript the segmentation workflow was applied to the largest available Region of Interest (ROI) that was clear of artefacts arising from air inclusions, with a volume of 382 $\times$ 355$\times$ 90 $\mu$m$^3$ (ROI 1).

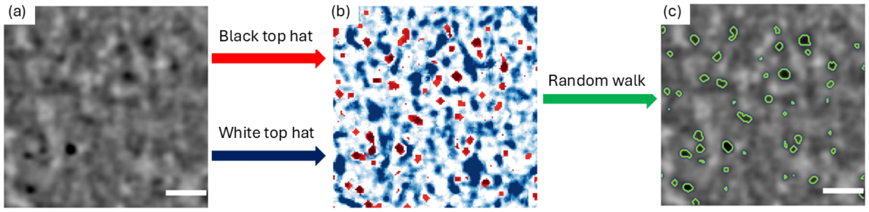


Figure 1: Automatic segmentation workflow. (a) Phase image. Scale bar 20 $\mu$m. (b) Image obtained by applying white top hat (blue) and black top hat (red) filters to the image of panel (a). (b) is used as seed for a random walk algorithm leading to the segmentation of cells' nuclei. Contours of segmented nuclei are shown in panel (c). Scale bars 20 $\mu$m.

To validate the nuclear segmentation workflow, three additional artefact-free ROIs were identified and the segmentation workflow applied. The volumes of the additional ROIs are 150 $\times$ 150$\times$ 75 $\mu$m$^3$ (ROI 2) and 150 $\times$ 150$\times$ 110 $\mu$m$^3$ (ROIs 3 and 4). Electron density, major and minor axes length, Feret diameter, eccentricity and volume were quantified for all segmented nuclei, with results shown in Figure 2. A good agreement across the morphological parameters is visible across the four ROIs. Some differences, however, can be found for the electron density distributions. This might be due to anatomical or pathological variations within the specimens. Indeed, Eckermann et al. [2] demonstrated a statistically significant difference in the nuclear electron density between healthy and pathological brain tissue.

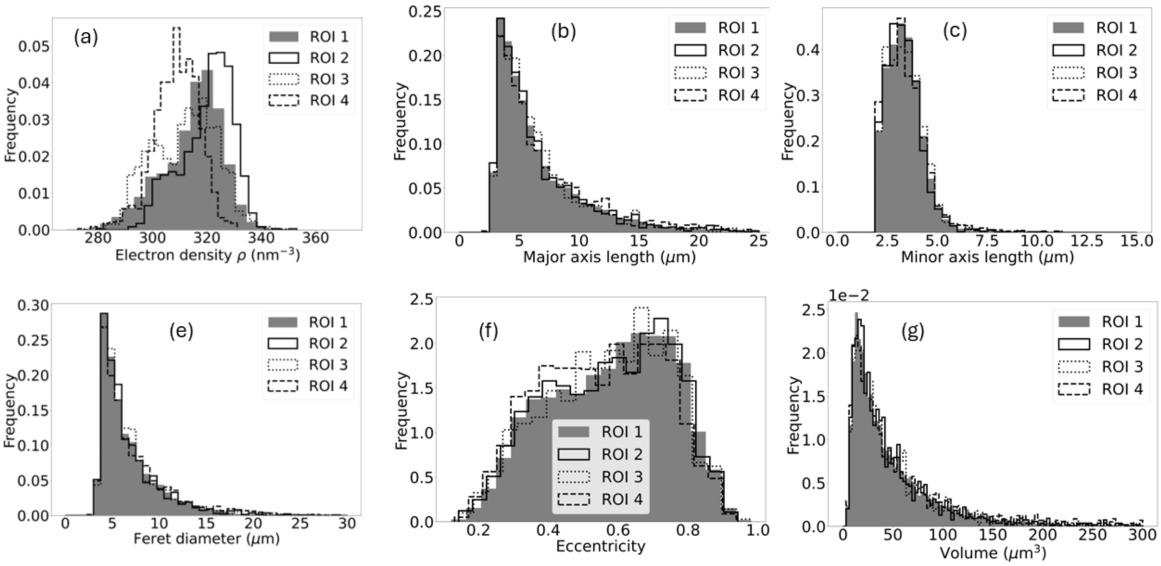


Figure 2: Nuclear morphological parameters for different ROIs. Electron density (a), major (b) and minor (c) axes length, Feret diameter (d), eccentricity (e) and volume (f). ROI 1 (solid grey) corresponds to the dataset presented in the main manuscript.

### Quantitativeness of the electron density maps

As detailed in the Methods section, the X-ray microscope enables retrieval of the refractive index decrement, $\delta$, from measured refraction data (see Equation 2), where $n = 1 - \delta + i\beta$ represents the complex refractive index. Furthermore, Equation 4 establishes that $\delta$ is directly proportional to the electron density, $\rho$. The quantitativeness of the measured $\delta$ maps is enabled by the quasi-monochromaticity of the X-ray source used. Here, we validate the quantitative accuracy of the electron density maps generated by our method. To do so, we performed tomographic imaging of two reference samples: a 150 $\mu$m diameter nylon wire and a 180 $\mu$m diameter polybutylene terephthalate (PBT) wire. A tomographic scan was conducted over a full 360° in increments of 0.5° increments, with the sample translated (dithered) in 2.5 $\mu$m steps. The refraction signal was retrieved, integrated, and used to reconstruct $\delta$ maps, which were subsequently converted into electron density values ($\rho$) using Equation 4. Figure 3(a) displays a reconstructed slice containing both test objects. Corresponding histograms of the measured $\rho$ for two regions of interest (ROIs), marked by blue and red circles in panel (a), are shown in panel (b). The measured electron density for the nylon wire was $\rho$=290.6 nm$^{-3}$, with standard deviation $\sigma$=18.0 nm$^{-3}$ and standard error $s$=0.3 nm$^{-3}$. The expected value, assuming the following chemical composition for the wire $(C_6H_{11}NO)_n$, is 280 nm$^{-3}$. The measured electron density for the PBT wire was $\rho$=334.1 nm$^{-3}$, with standard deviation $\sigma$=13.8 nm$^{-3}$ and standard error $s$=0.3 nm$^{-3}$. The expected value, assuming the following chemical composition for the wire $(C_{12}H_{12}O_4)n$, is 312 nm$^{-3}$. Theoretical electron density values were obtained from tabulated $\delta$ values [3].

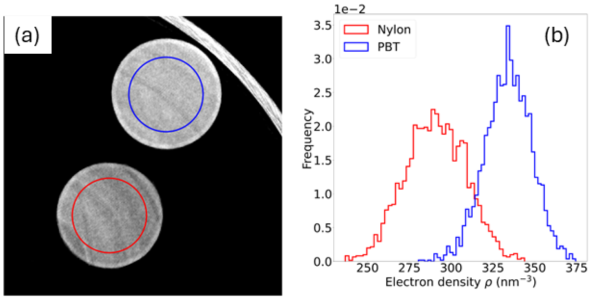


Figure 3: a) Electron density map for nylon and a PBT wires and corresponding histograms for electron density (b).

### Virtual histology: converting electron density maps to H&E histology volumes

In Figure 3 of the main manuscript, we demonstrated how a machine learning (ML) approach based on a Generative Adversarial Network (GAN) enabled the translation of measured electron density maps into three-dimensional H&E-like volumes. Owing to the lack of paired data between the two domains, we employed a CycleGAN [4] architecture, an approach widely used in histopathology for stain transfer [5, 6].

The network, designed to translate images from domain A (phase CT) to domain B (H&E histology), consists of two generators G_A (A→B) and G_B (B→A) along with two discriminators D_A and D_B. In the CycleGAN framework, the generators perform image translation between domains, while the discriminators aim to differentiate real images from those generated in each domain. The generators and discriminators are trained concurrently but independently. The training process involves several loss functions: the generative loss (G_x_Loss), the discriminative loss (D_x_Loss) and the cycle consistency loss (Cycle_x_Loss), where x denotes either domain A or B. The generative loss assesses the quality of image translation between domains, while the discriminative loss quantifies the ability of the discriminator to distinguish real from synthetic images. Through adversarial training, the model iteratively refines the generators to produce images that are increasingly realistic and challenging for the discriminators to classify. Additionally, the cycle consistency loss, proposed in [4], penalizes discrepancies between the original and reconstructed images, thereby reinforcing the fidelity of domain translation.

The training and testing datasets consisted of 1080 and 1322 patches for phase CT and histology, respectively. Histological images were obtained from the same sample, although not paired with the CT dataset. Each patch (224×224 pixels) was extracted by resampling the original data with a sliding window strategy and a stride of 100 pixels. Phase CT images were pre-processed to artificially increase the contrast of the nuclei using the segmentation labels. A value corresponding to 80% of the maximum was assigned to pixels corresponding to segmented nuclei. For the histology data, the slide background was masked, i.e. areas of the histological slide not containing tissue were excluded. The he resampled patches included the background for less than 0.1% of the total pixels. The network was trained for 200 epochs. A learning rate of 0.0002 with a linear decay to 0 after 100 epochs was applied, to prevent over-fitting. The instance normalisation layer was used in the CycleGAN to normalise each patch individually before training. After completing the training of CycleGAN, only the generators (G_A and G_B) were utilized to perform image translation between the two domains, whereas the discriminators were excluded during the inference phase.

To evaluate the performance of the CycleGAN model, we employed a 5-fold cross-validation strategy. An 80:20 split was applied to divide the data into training and validation datasets. For each fold, patches were randomly assigned either to the training or validation sets. The model was trained *ex novo* and tested, with this process repeated five times (once for each fold) to ensure all data were used for validation. No paired samples were shared across training and validation sets to preserve domain independence. The same network architecture and training hyperparameters were used across all folds.

To quantitatively assess the correspondence between the generated H&E-like volumes and the reference CT images, we calculated the Dice Similarity Coefficient (DSC) on binary masks obtained from the validation datasets after training at each fold. The DSC quantifies the spatial overlap between predicted and reference regions, with values closer to 1 indicating higher agreement. DSC were computed on a per-slice basis and averaged across all validation samples. Binary masks were obtained by segmenting nuclei in both CT and generated histology images. DSCs obtained for each of the folds are reported in Table 1. Among them, fold 1 achieved the highest DSC of 0.653 and was therefore selected as representing the best correspondence between the two domains.

Figure 5 illustrates the model's ability to accurately reproduce nuclear structures in the generated histology. Panels (a–c) and (d–f) compare the original CT images and the synthetic histology, respectively, while panels (g–i) and (j–l) display the corresponding nuclear segmentations. The comparison shows that the positions and shapes of nuclei are generally well preserved from CT to histology; however, the nuclei in the generated images tend to appear enlarged. This is further highlighted in panels (m–o), which show the differences between the two sets of labels. These discrepancies are mostly localized around the nuclear boundaries and can be attributed to the difference in slice thickness between the histological sections (4 $\mu$m) used for training and the CT scans (0.75 $\mu$m). While this represents a key limitation of the virtual histology approach presented in this study, future work could mitigate this issue by using thinner histological sections—potentially as thin as 1-$\mu$m—which are expected to reduce such discrepancies.

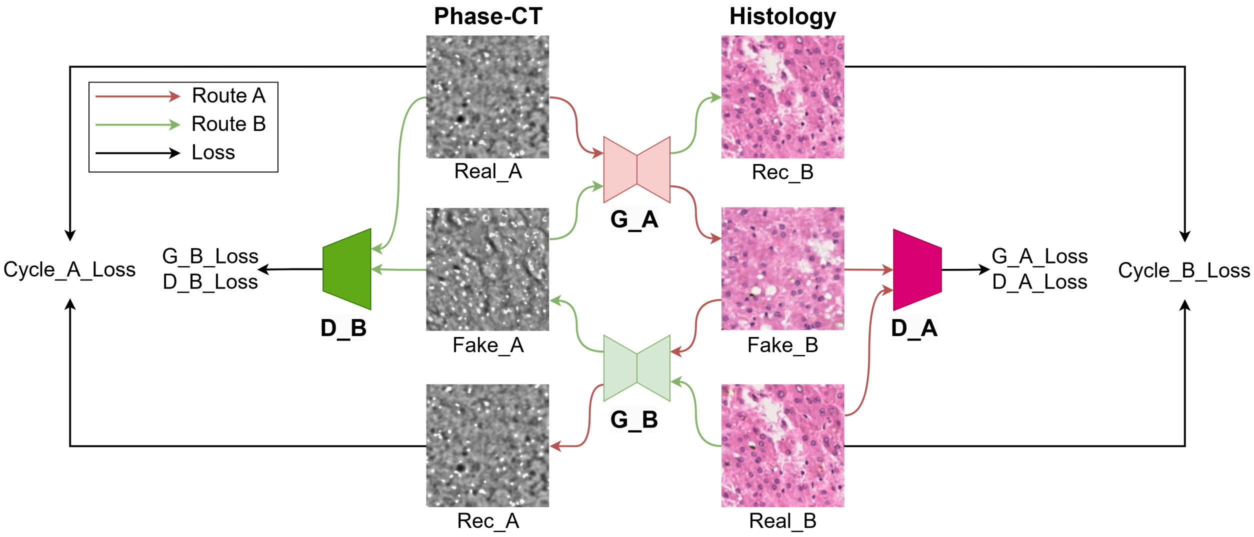


Figure 4: A schematic representation of the cycleGAN architecture transferring style A (phase CT) to style B (H&E histology). The network comprises two generators (G_A, G_B) and two discriminators (D_A, D_B). Loss functions of individual components as well as over the cycles are indicated.

| Fold | 0 | 1 | 2 | 3 | 4 |
|---|---|---|---|---|---|
| **Dice** | 0.641 | 0.653 | 0.611 | 0.527 | 0.635 |

Table 1: Dice score measured on the test dataset for each fold training.

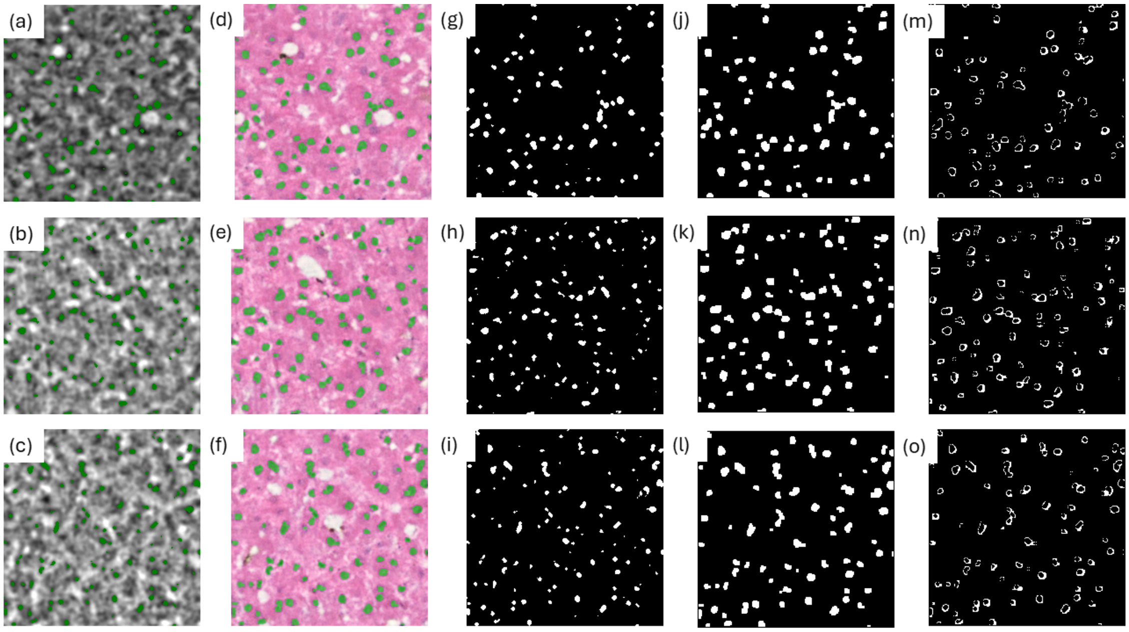


Figure 5: Validation of the style transfer. Phase CT (a-c) and corresponding generated histology (d-f) with overlaid labels segmented nuclei (in green). Segmentation labels for CT (g-i) and generated images (j-l). Difference between generated and CT labels (m-o). All images are $168 \times 168$ $\mu$m$^2$.

### Opportunity for reducing exposure time: the cycloidal approach

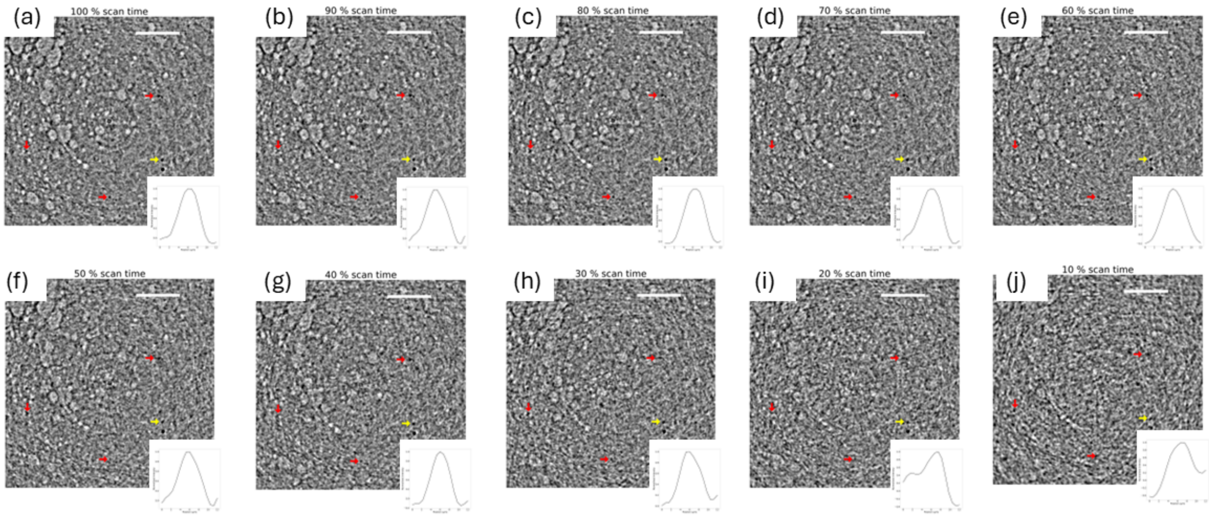


Figure 6: Phase CT slice obtained using an increasing reduced number of dithering steps, leading to an exposure time from 100% of the original dataset (a) to 10% of it (j). Red arrows highlight a few reference nuclei. Yellow arrows identify a nucleus used for plotting its intensity profile, shown as an inset. Scale bars 100 $\mu$m.

The results presented in this work are achieved through the use of attenuation masks composed of alternated transmitting slits and absorbing septa, which produce a structured illumination pattern of intensity peaks and troughs (see Figure 1(a) of the main manuscript). As a result, at any given acquisition step, only a subset of the sample is probed. To ensure complete sample coverage, a *dithering* process is employed, whereby the sample is incrementally stepped perpendicular to the direction of the transmitting apertures (see Figure 1 in the main manuscript), over a total distance equal to the mask's period. While guaranteeing phase sensitivity and the key advantage of aperture-driven resolution [7], this process entails stepping the sample at each acquisition angle of a tomogram, resulting in longer acquisition times.

To overcome this limitation, a cycloidal scanning approach has been proposed [8] where only a single step is acquired at each projection angle and the resulting sparse sinogram is interpolated. To evaluate the suitability of the cycloidal scan approach in the context of X-ray microscopy, the tomographic data of the liver tissue presented in the main manuscript have been undersampled according to the cycloidal scheme and reconstructed to simulate different scenarios of exposure time from 100% of the original dataset down to 10%. Specifically, sparse sinograms have been obtained from the original dataset by randomly sampling a number of dithering steps $N_d \in [1, 10[$ with $N_d = 10$ corresponding to the full dataset. The resulting sparse sinograms have been interpolated using a *Nearest-neighbour* interpolator, followed by phase integration and tomographic reconstruction, as detailed in the Methods section. Reconstructed tomographic slices are reported in Figure 6. Red arrows indicate specific features of interest, namely cell nuclei, while a yellow arrow marks a representative nucleus used for quantifying an intensity profile. The corresponding intensity profile is displayed as an inset in each panel. Although an increase in noise can be seen as exposure time decreases, even with a reduction in exposure time of up to 50% (i.e. $N_d = 5$, panel (f)), cell nuclei remain clearly resolvable, and no significant alterations are observed in the corresponding intensity profiles. However, for shorter scan durations, noise becomes the dominant factor and a decrease in resolution is observable, leading to a loss of resolution in high-frequency features such as nuclei. It is to be noted that all the tomograms in Figure 6 consist of the same number of angular projections, due to data availability. It has been shown [9] that for the cycloidal scheme to preserve aperture-driven resolution, specific sampling conditions, including angular, have to be met.
